# Medications that Regulate Gastrointestinal Transit Influence Inpatient Blood Glucose

**DOI:** 10.1101/2024.07.31.24311287

**Authors:** Amanda Momenzadeh, Caleb Cranney, So Yung Choi, Catherine Bresee, Mourad Tighiouart, Roma Gianchandani, Joshua Pevnick, Jason H. Moore, Jesse G. Meyer

## Abstract

**Objective:** A multitude of factors affect a hospitalized individual’s blood glucose (BG), making BG difficult to predict and manage. Beyond medications well established to alter BG, such as beta-blockers, there are likely many medications with undiscovered effects on BG variability. Identification of these medications and the strength and timing of these relationships has potential to improve glycemic management and patient safety.

**Materials and Methods:** EHR data from 103,871 inpatient encounters over 8 years within a large, urban health system was used to extract over 500 medications, laboratory measurements, and clinical predictors of BG. Feature selection was performed using an optimized Lasso model with repeated 5-fold cross-validation on the 80% training set, followed by a linear mixed regression model to evaluate statistical significance. Significant medication predictors were then evaluated for novelty against a comprehensive adverse drug event database.

**Results:** We found 29 statistically significant features associated with BG; 24 were medications including 10 medications not previously documented to alter BG. The remaining five factors were Black/African American race, history of type 2 diabetes mellitus, prior BG (mean and last) and creatinine.

**Discussion:** The unexpected medications, including several agents involved in gastrointestinal motility, found to affect BG were supported by available studies. This study may bring to light medications to use with caution in individuals with hyper- or hypoglycemia. Further investigation of these potential candidates is needed to enhance clinical utility of these findings.

**Conclusion:** This study uniquely identifies medications involved in gastrointestinal transit to be predictors of BG that may not well established and recognized in clinical practice.

## Background and Significance

Individuals with diabetes are at a significantly increased risk of hospital admission, with rates ranging from two to six times higher than those without diabetes^1–4^. Uncontrolled blood glucose (BG) in the hospital is associated with increased morbidity and mortality.^5^ Inpatient hypoglycemia, or BG<70mg/dL^6,7^ that occurs during hospitalization, has been reported in 20% of hospitalized insulin-treated patients^8^ and is the most common adverse event associated with inpatient treatment of diabetes^5,9,10^. Hypoglycemia can cause troublesome acute symptoms such as confusion, impaired vision and seizures, but can also increase the risk of chronic complications, length of stay and all-cause mortality^11–17^. Inpatient hyperglycemia, defined as a BG>140mg/dL in hospitalized patients^5^, can cause acute (e.g., diabetic ketoacidosis) and long term (e.g. retinopathy, nephropathy, cardiovascular disease) complications, both of which are associated with an increased risk of mortality^18^.

The dangers associated with dysglycemia highlight the importance of researching mechanisms involved in glucose homeostasis. Numerous pharmacologic agents are well established to cause hyperglycemia, including steroids and beta-blockers^19^, through a variety of mechanisms such as impaired insulin secretion or direct effects on beta cell proliferation^20^. However, many of the mechanisms underlying glucose alterations due to drugs are not well understood^20^. Drug package inserts may not be a reliable source for adverse events as they may present data from studies limited by small sample size and incidental findings^21^. Confounding is present between patient risk factors and BG^22^. For example, a drug administered to an individual with diabetes may be reported to cause hyperglycemia, but this correlation may be due to the disease and not actually the drug^21^.The management of BG in hospitals is further complicated by the dynamic nature of medication prescribing and patients’ unpredictable responses to these medication changes^23^.

Approved medications may have unexpected effects. In some cases, positive developments have been made in drug re-purposing research. For example, bumetanide, a diuretic, has been suggested as a potential therapy for Alzheimer’s disease^24^. Attempts have been made to identify drugs that may be repurposed for diabetes treatment. A computational pipeline using a large type 2 diabetes mellitus (T2DM) genome-wide association study combined with outpatient electronic health records (EHR) found angiotensin-converting enzyme inhibitors and calcium channel blockers to affect BG^25^. Another study created an open source repository with drug targets and genes specific to diabetes. Investigators screened around 1500 drugs on 20 proteins and identified five drugs with potential for repurposing to treat diabetes^26^. A data-guided approach and the wealth of information stored in the EHR may provide an initial step in uncovering drugs not previously established to cause BG alterations.

To address this challenge, we extracted a large, comprehensive EHR dataset including hospitalized patients who had received at least one anti-diabetic medication. The 500 most frequently prescribed inpatient medications were combined with relevant past medical history, demographics, social history, and labs available in the EHR. A least absolute shrinkage and selection operator (Lasso) linear regression model optimized using a training set allowed for filtering those initial variables using the highest model coefficients^27^. A linear mixed model (LMM) was fit and tested on the selected features from Lasso. LMM accommodates inter-patient variability and allows the interpretability through statistical significance testing that is lacking with machine learning models. This study aimed to use a computationally driven strategy and a large dataset to reveal associations between medications and point of care blood glucose (POC BG). To our knowledge, this is the first study to evaluate the predictive performance of several hundred medications without filtering based on prior knowledge of medications known to effect BG.

## Materials and Methods

### Data Source

Available retrospective EHR data was extracted for patients admitted to the hospital between May 2014 and September 2022 within the Cedars-Sinai Health System, which includes a tertiary medical center, Cedars Sinai Medical Center (CSMC), and community hospitals, Marina Del Rey Hospital and Huntington Hospital. Inclusion criteria consisted of adult inpatients (>/= 18 years old) who received at least one anti-hyperglycemic medication during their admission. Encounters to Labor & Delivery, Emergency Department, or with a length of stay less than 24 hours were excluded. The following EHR datasets were extracted for each encounter: past medical history, demographics, social history, labs, and inpatient medications. Cedars-Sinai IRB approval was obtained for this study.

### Model Design

Model inputs were static and time-dependent variables (**Figure 1A**). Static variables, or baseline characteristics that do not change during an inpatient stay, included demographic data, past medical history, and social history, and were collected from a single point during the admission (**Table 1**). Time-dependent variables, or variables that are changing during an admission, included labs and medications and were collected during a 12-, 24- or 48-hour lookback window preceding a prediction horizon. Given POC BG is checked typically four to six times per day in the hospital, a 4-hour prediction horizon was used to mimic a realistic next POC BG check. An output POC BG was selected for each encounter if there was another POC BG recorded between 16 and 4 hours prior to it to ensure maximal number of patients included in the analysis.

**Table 1.** EHR datasets, number of variables per dataset, and variables extracted from each dataset that were included in the study. The combination of numbers and letters in parentheses in the past medical history row are international classification of diseases, tenth revision (ICD-10) codes.

|  | EHR Dataset | Number of variables | Model Variables |
| --- | --- | --- | --- |
| Static | Past Medical History (ICD-10 code) | 12 | Liver failure (K72), chronic kidney disease (N18), type 1 diabetes mellitus (E10), type 2 diabetes mellitus (E11), malignancy (C80.1), anemia (D64.9), congestive heart failure (I50), hypothyroidism (E03.0), hyperthyroidism (E05), pregnancy (Z33.1), hypoglycemia (E16.2), pain (R52) |
|  | Demographic | 14 | Ethnic group, sex, race, smoking tobacco last use status, intravenous drug last use status, alcohol last status, admission diagnoses: congestive heart failure, sepsis, gastrointestinal bleed, nausea/vomiting, altered mental status, acute kidney injury, end stage renal disease +/- dialysis, pain |
|  | Social History | 4 | Age, body mass index, hospital location upon admission (intensive care unit or operating room) |
| Time-dependent | Laboratory | 5 | Mean POC BG, minimum POC BG, last POC BG, maximum POC BG, mean creatinine |
|  | Inpatient Medications | 516 | Top 500 most administered medications, commonly prescribed anti-hyperglycemic medications, TPN |

**Figure 1.**
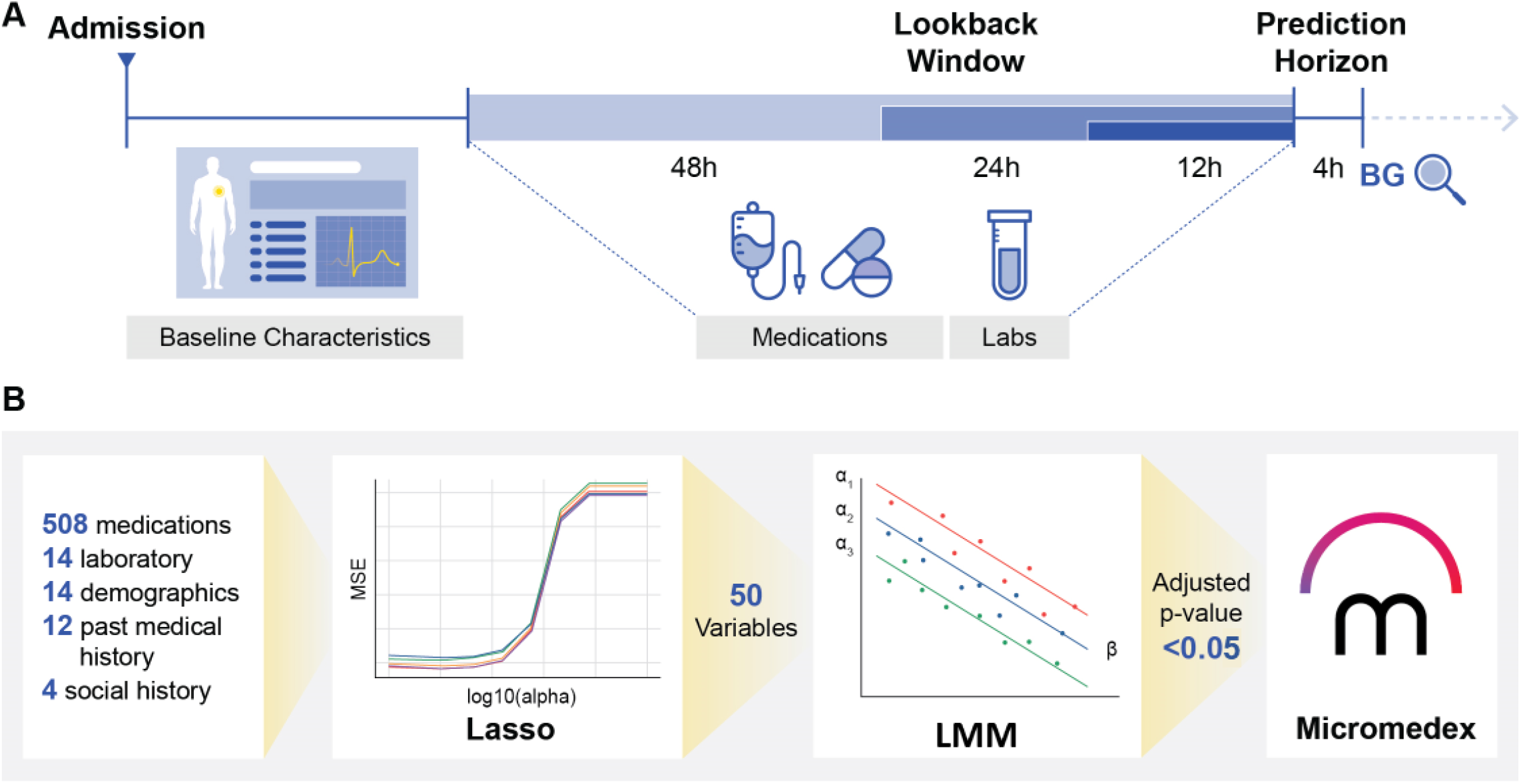
Project Overview. **(A)** Collection of model inputs (baseline characteristics, medications, and labs) and output POC BG variables over course of patient admission. **(B)** Data analysis pipeline demonstrating extraction of most important variables through Lasso, assessment of statistical significance using LMM and evaluation of novelty of medication-induced BG variability using Micromedex.

### Model Inputs

Medications were filtered to the 500 most commonly administered out of the total list of 5,267 medications. To ensure inclusion of common anti-hyperglycemic medications in the model, all subcutaneous (SQ) insulins (lispro, regular, glargine and NPH/Regular 70-30), intravenous (IV) regular insulin and all dosage forms of commonly prescribed oral anti-hyperglycemic medications (sitagliptin, glimepiride, glipizide, glipizide XR, glyburide and pioglitazone, nateglinide) were added to the top 500 if they had not already been included in the top 500 list. Total parenteral nutrition (TPN) was also added to the list of medications. All irrigation, flush, and iohexol orders were excluded due to inconsistency in recording of administered doses. This resulted in 516 unique medications. POC BG and creatinine were selected as input labs as they were recorded reliably during lookback windows and are shown to be predictive of BG alterations^9,23^. Past medical history and admission diagnoses were filtered for co-morbidities previously described to be predictive of BG variability^28–31^. Past medical history was obtained from international classification of diseases, tenth revision (ICD-10) codes, which were obtained from all available sources in the EHR (e.g., physician billing, problem list). **Table 1** lists all static and time-dependent variables selected for downstream analysis.

### Data Pre-processing

Missing data was dealt with in one of two ways. Lab values and medication doses that were missing in the dataset were removed. Categorical variables that were recorded as declined, missing or unknown were retained and re-coded as missing. Medications that were not administered during a lookback window were assigned a zero value. These five parent datasets (medications, laboratory, demographics, social history, and past medical history) were merged on shared unique patient encounter numbers, resulting in 87,256 encounters (49,095 patients) using a 48-hour lookback, 78,001 encounters (45,819 patients) using a 24-hour lookback, and 53,576 encounters (35,459 patients) using a 12-hour lookback.

Categorical variables were dichotomized. The sum of all doses for each medication was calculated during the lookback window and were used as inputs to the model. Mean, minimum, maximum and last value of POC BG and mean of creatinine were calculated over each lookback window and used as inputs to the models.

### Statistical Analysis

Feature selection was performed due to lack of convergence of the LMM when attempting to use all features. Eighty percent of the encounters were used for training and the remaining 20% for testing. Feature selection was performed using the 80% training set, which was further split into 80% train and 20% validation sets to avoid data leak into the test set during feature selection. Lasso regression, a linear model with a regularization term, was selected to perform feature selection since we expected that out of the many predictors to the model, only a subset of predictors had a non-zero effect. The regularization parameter of Lasso drives less informative variables to have coefficients of zero and helps identify features most predictive of the outcome^27^. Lasso models were constructed using input variables collected over lookback windows of 12h, 24h, and 48h. Lasso 5-fold cross-validation determined the best regularization parameter (alpha) among one hundred alpha values between 0.0001 to 10,000 based on the lowest mean squared error (MSE) mean over each fold. Using this best alpha, the trained model was then used to predict BG in the validation set and performance was evaluated by comparing the predicted BG with the true value using R2 (coefficient of determination) and root mean squared error (RMSE).

To ensure robustness and reproducibility in coefficient ranking, the Lasso model with the optimized alpha was run using repeated k-fold cross-validation with 5 splits, 5 repeats and a maximum iteration limit of 10,000 on the training data. The mean absolute value of the extracted coefficients (25 total coefficients per variable) was then calculated for each variable and sorted to identify the 50 variables with the highest mean absolute coefficient values.

The top 50 ranked predictors of BG from Lasso were inputted to a multivariate LMM to assess their statistical significance.^22^ Due to repeated encounters present per patient, we used LMM to predict an output BG. The random effects’ covariance structure was the variance component’s structure, where each group has its own random intercept. Variables with a Benjamini-Hochberg (B-H) adjusted p value <0.05 were considered to have statistically significant associations with BG. While not feasible to include every potential interaction term, we re-fit the LMM with clinically relevant interaction terms including history of congestive heart failure, acute pain, and whether the patient was admitted to the peri-operative setting. Variables that retained statistical significance using 12, 24, and 48h lookback windows were compared to understand differences in medications predictive of BG over different lookback time courses. The ‘Adverse Effects In-Depth Answers’ section of Micromedex, a comprehensive evidence-based resource used by clinicians^32^, was reviewed for documentation of BG alteration properties of statistically significant medications. In addition to adverse events reported in package inserts and clinical trials, Micromedex includes up to date post-marketing surveillance data and case studies. Statistical analysis was performed in Python version 3.9.12 using the following packages (versions): sklearn (1.4.2), statsmodels (0.13.5), scipy (1.11.4), and seaborn (0.12.2).

## Results

### Descriptive Statistics

**Figure 1A** depicts the timeline over which the model input and output variables are collected during a patient admission. A POC BG was selected as the output for an encounter there was another POC BG recorded between 4 and 16h prior to it. Input variables that do not change over the course of the hospital stay (demographics, past medical history and social history) were collected at the time of admission. We then collected input variables that change during an admission (medications and labs) over 12, 24, and 48h lookback windows ending 4 hours before the output BG was collected. **Figure 1B** demonstrates the data analysis pipeline, beginning with several hundred variables across datasets, the top 50 variables after Lasso, and finally only those with a B-H adjusted p-value less than 0.05 from the LMM. These variables were evaluated for prior documentation of affecting BG in Micromedex.

Baseline characteristics of the encounters from patients across three hospitals in a large urban health system collected over 8 years (2014 to 2022) used for model training and evaluation is summarized in **Table 2**. The median encounter age was 68.8 years, 63.3% were Caucasian, 18.8% Black or African American, and Hispanic ethnic group comprised 19.3% of encounters. 44.5% of all encounters were female and the median body mass index (BMI) was 27.0 kg/m^2^. Most encounters had diabetes; 10.3% had type 1 diabetes mellitus (T1DM) and 70.3% had T2DM. Nearly half of encounters had chronic kidney disease (CKD; 41.3%) and 8.8% had hypoglycemia listed as a past medical history prior to the encounter. The median and inter-quartile range (IQR) of the mean input POC BGs collected for each encounter during the 48h lookback window were 154.8 mg/dL and 127.0-194.2 mg/dL, respectively. 7.6% of encounters were admitted to the ICU and 1.13% were admitted to perioperative setting.

**Table 2.**
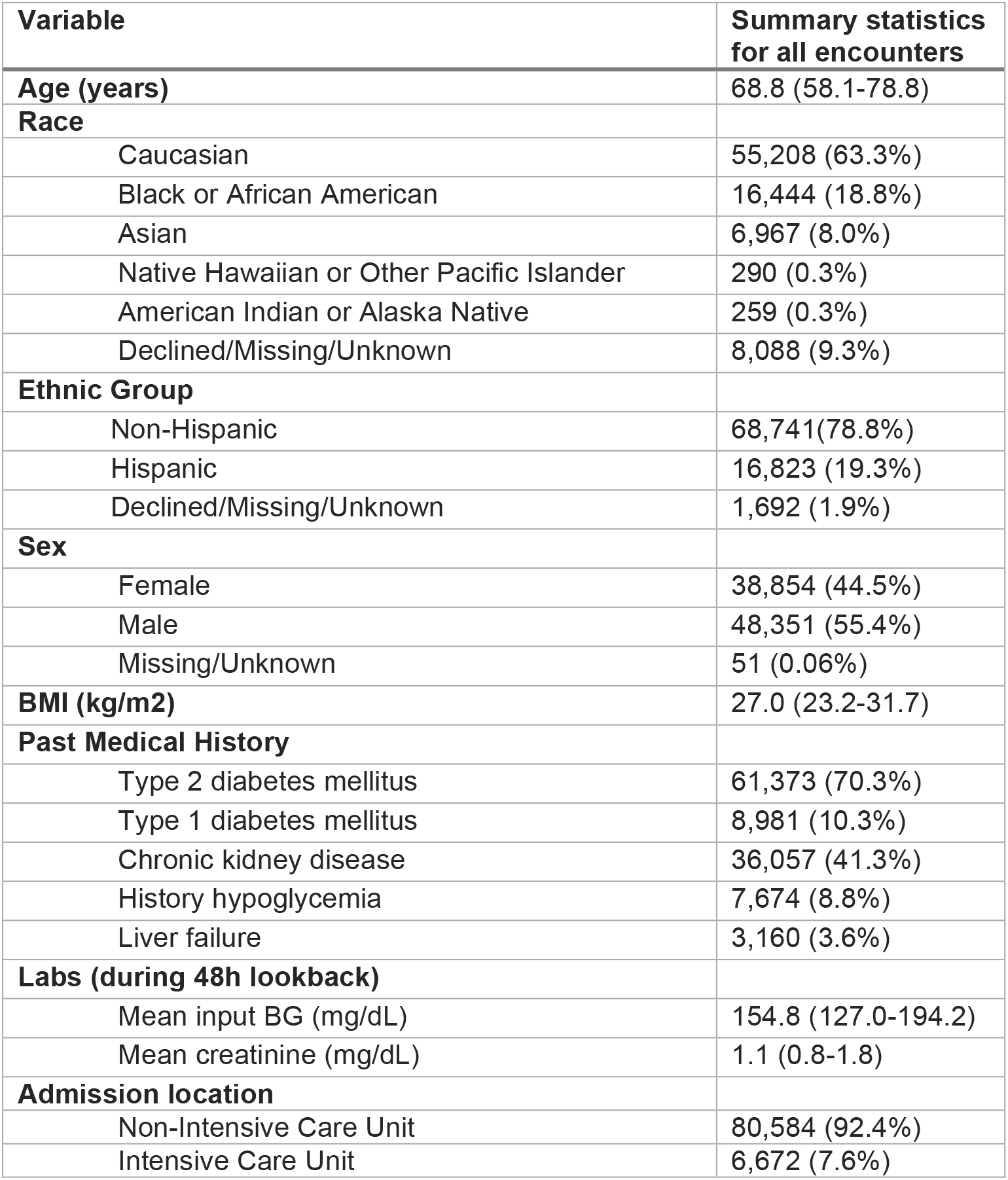
Summary statistics across patient encounters. Continuous variables are shown as median and IQR. Categorical variables are shown as counts and percent.

The top 10 administered medications out of the filtered 516 medications are displayed in **Figure 2A**. Three out of the ten medications were antihyperglycemics (insulin lispro SQ, insulin glargine SQ and insulin regular IV). Overlayed distributions of the input and output BGs are shown in **Figure 2B** with a Spearman correlation coefficient of 0.58, indicating that prior recorded BGs are correlated with the next BG.

**Figure 2.**
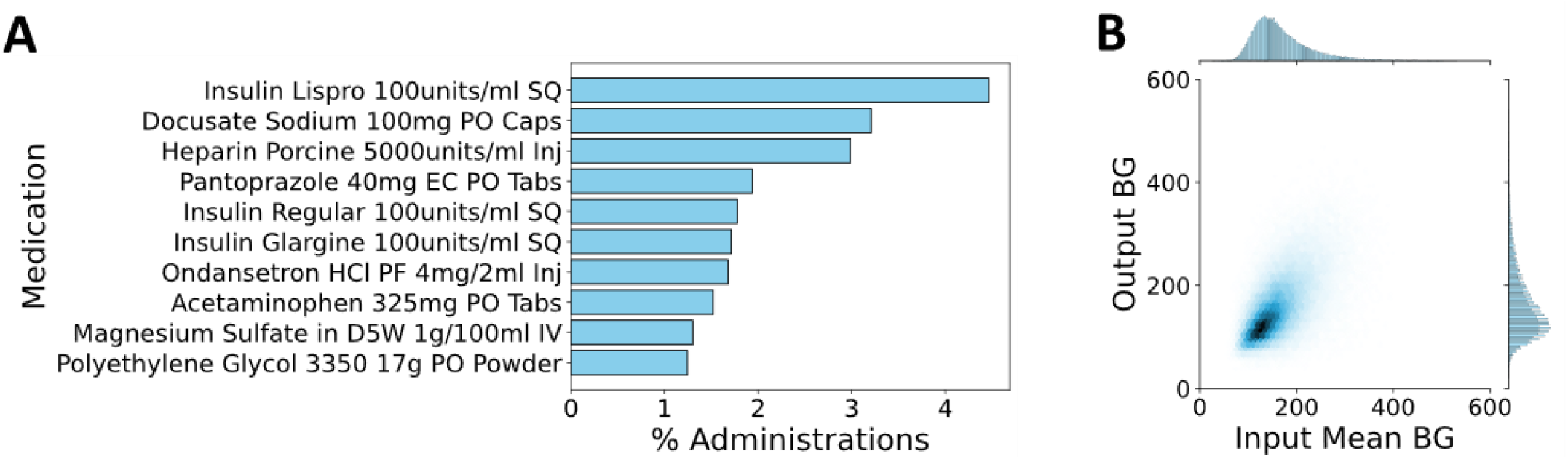
Overview of medications and BG data. **(A)** Top 10 most commonly administered medications. **(B)** Linear relationship between mean input BG over 48hr lookback window and output BG.

### Lasso-based feature selection

Performance for the Lasso model with a 48h lookback window was highest (R2=0.352) compared to 12h (R2=0.325) and 24h (R2=0.344) windows, and thus was used for the primary analysis presented. The 80% set (69,804 encounters) from the original dataset was further split to 80% training (55,843 encounters) and 20% validation sets (13,961 encounters). Lasso 5-fold cross-validation on the training set was used to fit a Lasso model. This model was then used to predict on the validation set, yielding R2 score 0.352 and RMSE 53. The alpha with the lowest mean MSE over the five folds using the training set was 0.36 (**Figure 3A**). Actual validation vs predicted output BG were linearly correlated (**Figure 3B)**. Lasso L1 regularization prioritized the most informative features while setting the coefficients of less important predictors to zero (**Figure 3C**). To ensure reproducibility in selection of top ranked coefficients, the Lasso model with the optimized alpha was run using repeated k-fold cross validation where the data was split into 5 folds and the process was repeated 5 times on the training data. Distribution of coefficient values calculated across folds/spits for the top 25 variables were stable (**Figure 3D**). The full list of 50 selected variables and their mean absolute value coefficient scores are shown in **STable 1**. The intercept of the Lasso model is the output value when all input variables are zero; the mean intercept across all folds was 52.

**Figure 3.**
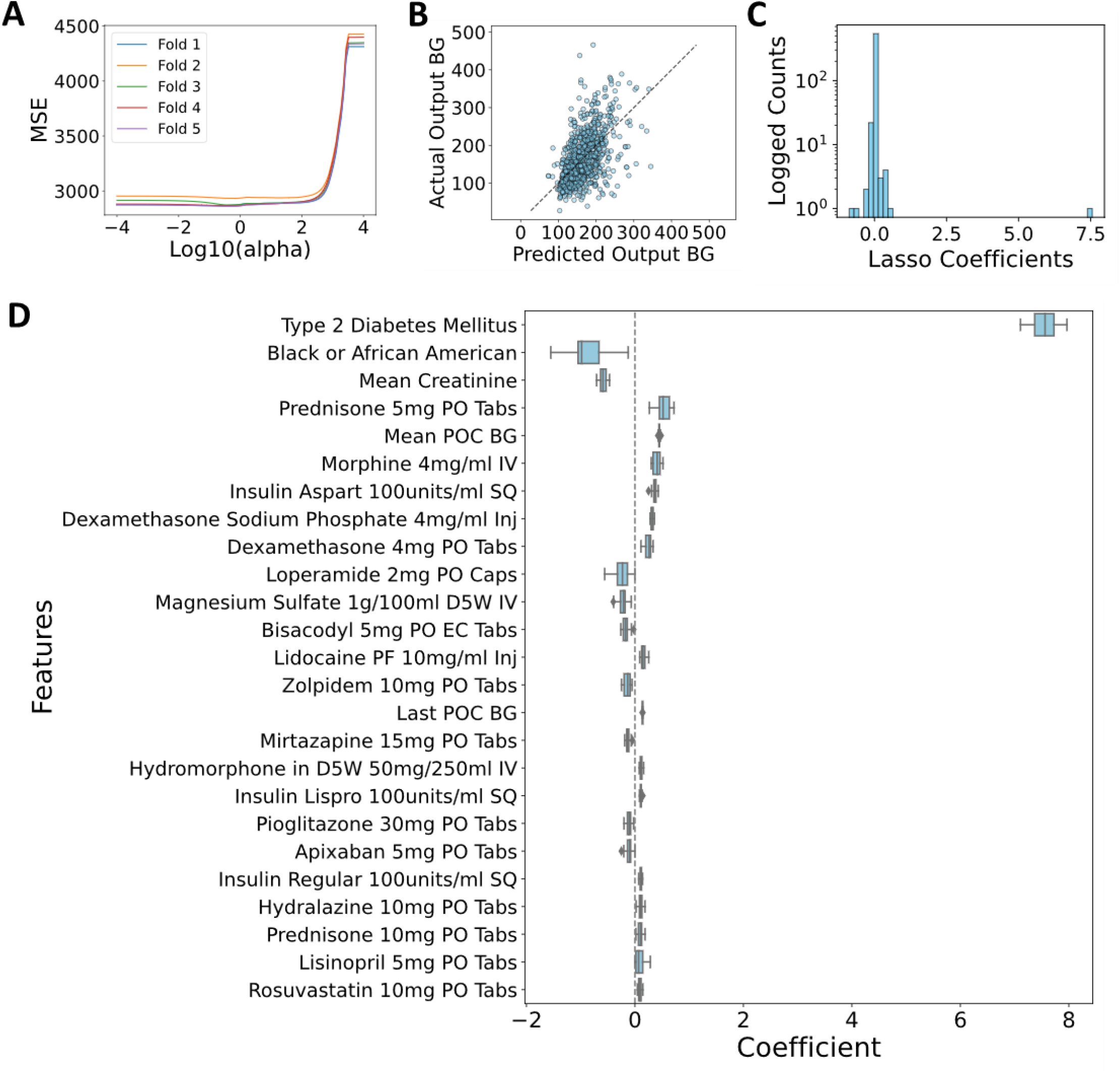
Overview of Lasso model optimization and feature selection results using 48h lookback window. **(A)** MSE vs log10(alpha) for each of 5 Lasso cross validation folds. (**B)** Actual validation vs Lasso-predicted output BG. **(C)** Variable coefficients assigned by Lasso. **(D)** Mean absolute coefficient score of top 25 Lasso variables.

### LMM and Micromedex

To address potential confounders, control for variability between each patient, and assess statistical significance of coefficients, a LMM was employed that expressed the output BG as a linear function of the 50 Lasso-selected input variables with a random intercept for each patient. The LMM was used to predict on the held-out 20% test set (i.e., not used for feature selection) with resultant performance metrics R2 0.342 and RMSE 53. The variance of the random intercepts, a representation of the between-group variability in output BG, was 57.6 (mg/dL)^2^. B-H adjusted p-values, coefficient, standard error, and 95% confidence interval (CI) for the 50 variables, intercept and group variable are shown in **STable 2**. Twenty-one variables that had been ranked in the top 50 variables by Lasso predictive of output BG did not meet the B-H adjusted p-value cut off 0.05. Of the remaining 29 variables (**Table 3**), 24 were medications and 5 were a combination of labs, demographics, and past medical history. More than half of the medications were documented in Micromedex as having a direct effect on BG, including insulins, glucocorticoids, lactulose, lisinopril, rosuvastatin and pioglitazone. All glucocorticoids, dexamethasone, methylprednisolone, and prednisone, were positively correlated with BG, as expected. The coefficient indicates the change in the output BG for a one unit increase in the input assuming all other factors influencing BG remain constant. For example, a coefficient of 0.78 for prednisone 5mg tablet indicates that, on average, taking an additional dose of prednisone is associated with an increase in BG by 3.9 mg/dL. Insulin lispro, aspart and regular SQ were positively correlated with a predicted BG measured at 4h while insulin glargine was negatively correlated with output BG. Mean input BG was positively correlated and had a stronger effect compared to last prior input BG. Mean creatinine was negatively correlated with BG. History of T2DM had a strong positive association with BG and was associated with an increase in BG by 10.66 mg/dL given all other variables are held constant.

**Table 3.**
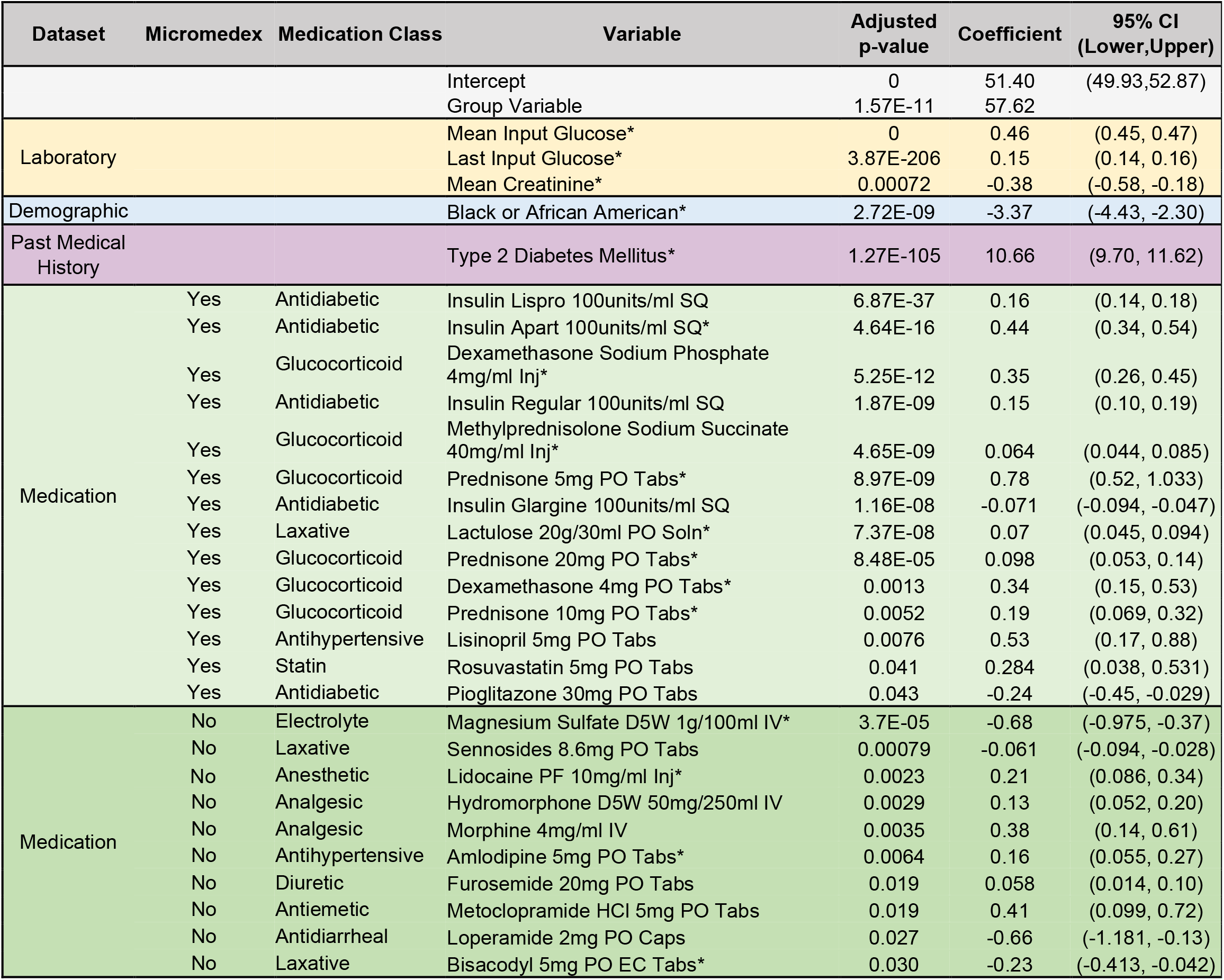
Twenty-nine variables with an adjusted p-value <0.05 according to LMM. EHR dataset, whether the medication was documented in Micromedex as having an effect on BG, the drug class, B-H adjusted p-value, coefficient, and 95% CI are listed. *denotes variable was significant across 12h, 24h, and 48h lookback windows.

Black/African American race was strongly negatively correlated, and was associated with a decrease in BG by 3.37mg/dL. Ten variables met the adjusted p-value cut-off of <0.05 and were not recorded as causing alterations in BG in Micromedex. Of those ten, medications negatively correlated with BG included magnesium sulfate, sennosides, loperamide and bisacodyl. Positively correlated were metoclopramide, lidocaine, hydromorphone, morphine, amlodipine, and furosemide.

The LMM was fit with the same 50 variables plus the inclusion of interaction terms to differentiate the effects of the underlying disease processes known to cause dysglycemia from the medication. Interactions assessed were known risk factors for hyperglycemia, including: (1) acute pain^3^ with morphine and hydromorphone, (2) history of congestive heart failure^34^ with furosemide, and (3) surgery^35^ with bisacodyl, lidocaine, loperamide, metoclopramide, and sennosides. No interaction term had an adjusted p-value less than 0.05, and each medication alone retained significance after inclusion of the eight interaction terms in the model. This indicates presence of the potential confounding factor did not differentiate the effect of the medication on the outcome (**STable 3**). **SFigure 1** allows visualization of the relationship between mean medication doses over the 48-hour lookback window and output BG across patient encounters.

Lastly, we observed differences in variables that were statistically significant based on the length of lookback window used (**Figure 4**). Adjusted p-value, coefficient, standard error, and 95% CI for 12h lookback window (**STable 4)** and 24h lookback window (**STable 5**) are presented in supplementary summary tables. Eleven variables of the significant variables using the 12h lookback window were unique to that window, five were unique to the 24h lookback window, and five were unique to the 48h lookback window. Trends were observed in varying statistical significance based on duration over which mean medication doses were collected. For example, pioglitazone, an antihyperglycemic with a half-life of 3-7 hours^36^, was significant only when a 48h lookback window was used while nateglinide, an antihyperglycemic with a half-life of 1.5h^37^, was significant only when a 12h lookback window was used.

**Figure 4.**
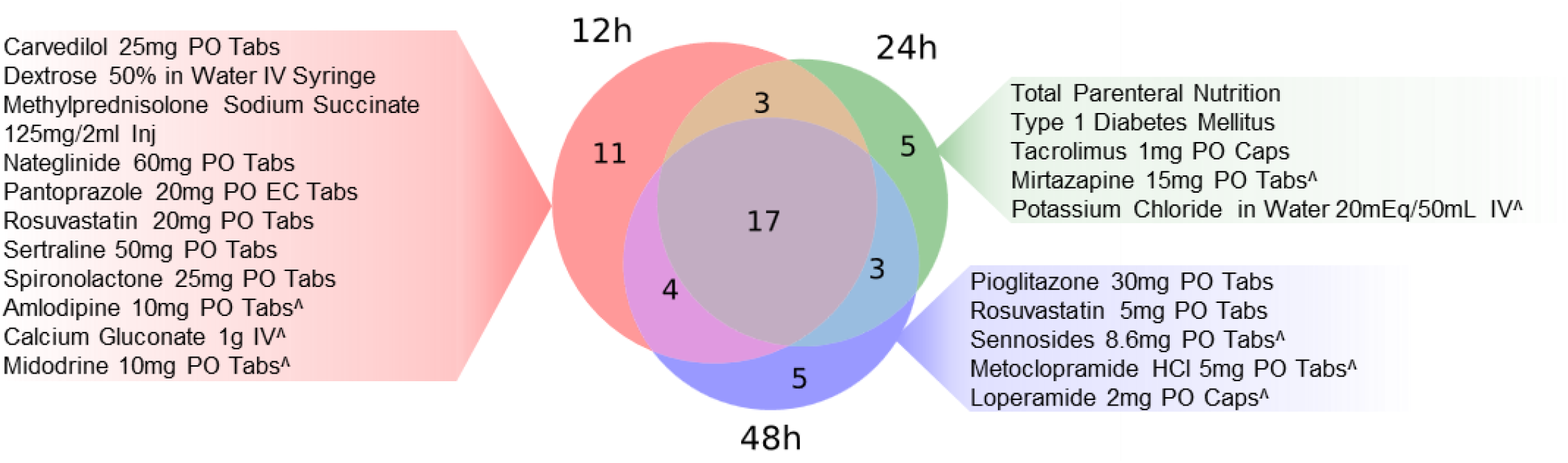
Significant variables that are unique to each lookback window. ^ denotes medication was not recorded in Micromedex as having the potential to affect BG.

## Discussion

To our knowledge, this is the first study that identifies medication predictors of BG alterations using a large EHR dataset. Mean and last input BG were positively correlated with output BG, and retained significance across lookback windows. Studies show that prior BG is important in predicting future BG^38^. Additionally, history of T2DM was strongly positively correlated with output BG across all three lookback windows. We found that Black/African American race was also strongly negatively correlated with output BG. Similar trends in BG among races have been demonstrated; after a glucose tolerance test, African Americans had a lower glucose area under the curve compared to Caucasians^39^. Several diabetes medications were statistically significant according to the LMM. We also observed several other non-diabetes medications well established to affect BG, including statins, carvedilol, dextrose 50% IV and TPN^20^. Interestingly, short and medium-acting insulins (regular SQ, insulin lispro, insulin aspart) were positively correlated with BG. These medications are often given prior to meals in response to elevated BG and have a peak response within 30-90 minutes^40^, duration of 3-4 hours^41^ and elimination half-life of approximately 1 hour^42^, so their BG lowering effect may be diminished at the four hour mark.

Several drugs involved in gastrointestinal (GI) motility, including sennosides, metoclopramide, loperamide, bisacodyl, lactulose and opioids, were statistically significant. The effect of gastric emptying on glycemic control is poorly understood. Colonic motility is slowed in the majority of individuals with diabetes,^43^ and retention of GI contents has been associated with hyperglycemia^44–46^. Studies indicate an initial hypoglycemia due to delayed carbohydrate delivery to the small intestine, followed by hyperglycemia;^44^ two hours after a meal, T1DM individuals with slowed gastric emptying had lower insulin requirements^47^, and use of a prokinetic agent led to increased insulin requirements^48^. We found that metoclopramide (a prokinetic agent) was associated with a higher BG at 4 hours while sennosides and bisacodyl were associated with a lower BG. Sennosides A, the active ingredient in sennosides, demonstrated the ability to stimulate intestinal cells to secrete GLP-1 and alter the gut microbiota^49^, leading to improved insulin sensitivity in mice^50,51^. Sennosides A was also shown to have similar potency as acarbose in alpha-glucoamylase inhibition.^52^ We found loperamide, which slows GI transit, to also lower BG. Loperamide may lower BG in insulin-deficient rats through increase of glucose transporter 4 (GLUT-4) expression and inhibition of hepatic gluconeogenesis^53^. Activation of mu-receptors by loperamide has also been linked to reduction of IL-6 induced insulin resistance^54^ and increase in insulin secretion by countering forkhead box protein O1 inhibition^55^. Lastly, we found morphine and hydromorphone to be associated with increases in BG. Opioids increase BG via direct effects on insulin secretion through opioid receptors in the pancreas and centrally, and activation of the sympathetic nervous system^56^. Opioids additionally reduce colonic transit time and lead to dysregulation of GI motility by binding the mu receptor and toll-like receptors in the GI tract.^57^ Overall, GI transit plays a large role in glycemic control, and we identified several digestive process regulating agents to be statistically important in predicting BG.

There was strong evidence in support of IV magnesium and diuretics. Magnesium intake has been shown to be inversely associated with T2DM risk^58,59^. Intravenous magnesium sulfate administration during surgery led to significantly lower BG and insulin requirements ^60,61^. A potential mechanism for improvement of insulin sensitivity is upregulation of GUT-4 gene expression, leading to increase in glucose utilization in peripheral tissues^62–64^. There is also strong evidence linking diuretics to insulin resistance with an association independent of confounders^65^.

Strengths of our study included an unbiased, rigorously test approach in finding medication predictors of BG. Rather than hand-picking known BG predictors, we allowed the Lasso model to perform feature selection.

Training data was used for feature selection to avoid test leak, and LMM performance was evaluated on a held-out test set. We also included both critically and non-critically ill patients as well as all types of insulin. We used a mixed effects model, which can handle repeated measures by accounting for the correlation between encounters of the same patient. Our model had comparable performance to a linear model that used a 24 hour moving average of inpatient BG measurements collected from EHRs to predict the next BG; R2 was 0.45 using all observations, and performance varied based on glycemic variability category (very high glycemic variability R2=0.14 to low glycemic variability R2=0.65)^38^. The focus of our study was to identify potentially undiscovered medication predictors of BG, rather development of a high-performing clinical model.

Limitations of our study included inability to control for the many confounders that may affect a patient’s BG. We did not include drug interactions that may be causing an alteration in BG. EHR data has several limitations, including missing data, erroneous entries, and lack of documentation of relevant variables. Diet and tube feed orders were not used due to lack of consistent recording, however TPN was included. There was a possibility for encounters from a single patient to be split across training and test sets. The dataset is large so even predictors that have small effect can be statistically significant. The linear regression method employed did not use time series data. However, a study using linear, cubist, random forest and K-nearest neighbors models to predict the next BG using previous BGs over various moving average and rolling regression windows found no difference in performance (R2 CI’s overlapped)^38^. Future directions include inclusion of more patient encounters to the model and assessment of its generalizability.

## Data Availability

Clinical data used for this study is not available.

## Code Availability

All JupyterLab notebooks can be found at xomicsdatascience/Medication-BG-alteration (github.com).

## Acknowledgments

We thank Edward Kowalewski and the Honest Enterprise Research Broker for EHR data extraction services and Somi Hwang for graphic design assistance.

## Funding and all other required statements

This research was supported by NIH National Center for Advancing Translational Science (NCATS), UCLA CTSI Grant Number UL1TR001881.

