## Supplementary Information for "Medications that Regulate Gastrointestinal Transit Influence Inpatient Blood Glucose"

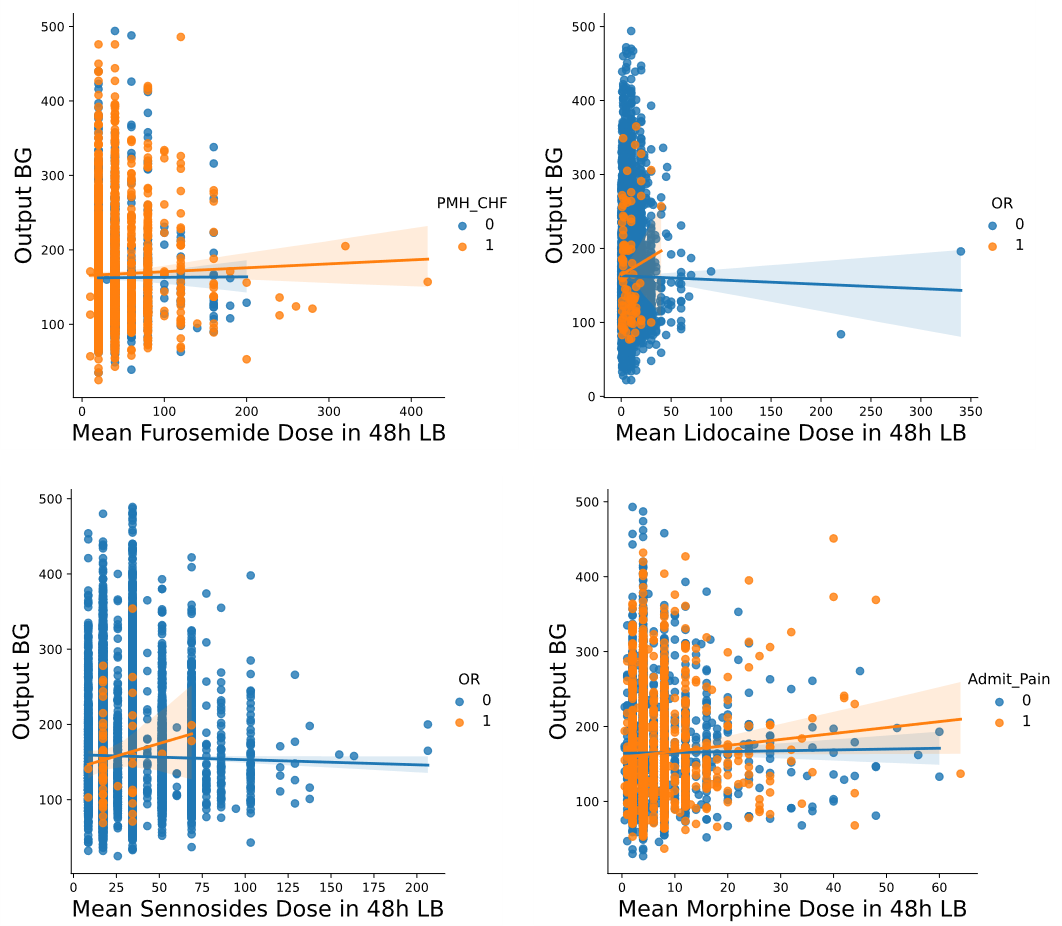

**Supplementary Figure 1.** Visual assessment of whether correlation varies between mean medication dose over a 48h lookback window and 4h output BG based on selected interaction term. Regression lines plotted indicate the general trend between each medication and the output value. Medications and interaction terms were: **(A)** furosemide 20mg oral tabs and history of congestive heart failure, **(B)** lidocaine 10mg/ml injection and admission to a peri-operative unit, **(C)** sennosides 8.6mg oral tabs and admission to a peri-operative unit, and **(D)** morphine 4mg/ml IV and admission for pain.
